# Causal Machine Learning for Comparative Effectiveness of GLP-1 RA versus SGLT2i in Heart Failure Using Real-World EHR Data

**DOI:** 10.64898/2026.04.06.26350259

**Authors:** Grace Y. Han, Andreas P. Kalogeropoulos, Zach Butzin-Dozier, Rachel Wong, Fusheng Wang

## Abstract

Clinicians lack precision medicine tools to estimate individualized treatment effects for patients with heart failure (HF). Causal machine learning leveraging electronic health records can estimate both average and individualized treatment effects, enabling estimation of treatment heterogeneity. Using Stony Brook University Hospital data, we compared the effectiveness of glucagon-like peptide-1 receptor agonists (GLP-1 RA) versus sodium-glucose cotransporter 2 inhibitors (SGLT2i) in patients with HF. Under a doubly robust framework, we found a stable population-average effect: GLP-1 RA was associated with a lower risk than SGLT2i for a 1-year composite outcome of all-cause mortality or HF-related hospitalization. Heterogeneity analyses provided limited evidence for individualized treatment selection, although subgroup tests identified loop diuretic use, body mass index, and estimated glomerular filtration rate as potential effect modifiers. While these models hold promise for translating observational data into actionable precision care, careful assessment of causal assumptions and rigorous validation are essential before clinical implementation.

## Introduction

Heart failure (HF) is a leading cause of morbidity, hospitalization, and death in the United States. Nearly 6.7 million U.S. adults live with HF, and the clinical burden continues to increase^1^. HF encompasses distinct clinical phenotypes with different underlying biology and therapeutic priorities^2^. In HF with reduced ejection fraction (HFrEF), outcomes are strongly influenced by maladaptive neurohormonal activation and cardiorenal congestion. In HF with preserved ejection fraction (HFpEF), the syndrome is more heterogeneous and often reflects a broader cardiometabolic profile. Guideline-directed medical therapy (GDMT) is central to HF management, although recommended therapies vary by phenotype. In HFrEF, GDMT emphasizes early initiation of multiple drug classes, including renin–angiotensin system inhibitors (RAASi), beta-blockers (BB), mineralocorticoid receptor antagonists (MRA), and sodium–glucose cotransporter-2 inhibitors (SGLT2i)^3^. In HFpEF, treatment is less uniform and generally focuses on symptom control and comorbidity management. As HF care becomes increasingly complex, clinicians must determine which therapies are most appropriate for individuals with diverse baseline characteristics.

Two medication classes are of particular interest for individualized treatment selection in HF: SGLT2i and glucagon-like peptide-1 receptor agonists (GLP-1 RA). Despite the well-established benefits of SGLT2i across HF phenotypes in the literature^4^, real-world adoption remains incomplete. In a registry study of 49,399 hospitalized HFrEF patients, discharge prescriptions were 68.2% for RAASi, 89.0% for BB, 41.0% for MRA, and only 20% for SGLT2i^5^. In parallel, large cardiovascular outcome trials of GLP-1 RA generally demonstrated significant reductions in major adverse cardiovascular events (MACE) in patients with diabetes or overweight/obese adults at risk for cardiovascular events, but modest effects on HF outcomes^6^. Their HF relevance appears more phenotype dependent. In patients with HFrEF, there are safety concerns from early trials with older GLP-1 RAs and adverse mechanistic signals^7^. In obesity-related HFpEF, semaglutide and tirzepatide improved HF symptoms in STEP-HFpEF and SUMMIT, respectively, supporting an emerging role for GLP-1 RA as a phenotype-targeted adjunct HF therapy^6,7^. However, uptake of GLP-1 RA in routine care is limited by access, tolerability, and workflow barriers.

Together, these developments highlight the need to better understand how to optimize therapy at the individual patient level. Patients with HF exhibit substantial heterogeneity in clinical phenotype, comorbidities, and metabolic risk, which may affect their response to different cardiometabolic therapies. Although SGLT2i is recommended, other agents such as GLP-1 RA may also improve outcomes in selected HF patients. However, clinicians currently lack patient-specific evidence to determine the optimal therapy for an individual and must instead rely primarily on population-level trial results and guideline recommendations.

Randomized clinical trials remain the gold standard for establishing population-level treatment effects, but they are costly, time-consuming, and underpowered to provide guidance on individualized treatment selection^8^.

Observational EHR data provide a scalable substrate for comparative effectiveness analyses in real-world populations, yet naïve comparisons remain associative because treatment selection is not randomized. Causal inference methods bridge this gap by defining a clear causal estimand and a target trial-like study design, then estimating treatment effects under explicit identifiability assumptions that move beyond associations^9,10^. Within this framework, traditional regression and propensity score approaches can be inadequate in complex EHR settings because they rely on restrictive modeling assumptions and sufficient covariate overlap^11^. Building on these ideas, causal machine learning (causal ML) uses flexible learners to better handle high-dimensional, nonlinear data, enabling the estimation of individualized treatment effects (ITE)^12^.

Causal ML methods have shown promise for comparing cardiometabolic therapies and predicting heterogeneity using real-world data. Wang et al. applied a causal forest-based method to compare SGLT2i and GLP-1 RA initiation in Medicare beneficiaries, using incident HF hospitalization as the outcome^13^. Cardoso et al. used Bayesian causal forest to compare SGLT2i versus GLP-1 RA in patients with type 2 diabetes mellitus (T2DM), with incident HF diagnosis as a long-term outcome^14^. However, in populations with established HF, treatment initiation is often influenced by illness severity, which induces confounding by indication and thereby complicates stable estimation of treatment effects. To address this gap, we used electronic health records (EHR) data from Stony Brook University Hospital (SBUH) to compare initiation of GLP-1 RA versus SGLT2i among patients with HF. We implemented an analytic causal ML framework using doubly robust methods to estimate both population-average and individualized treatment effects, while evaluating whether such heterogeneity signals were credible under real-world prescribing.

## Methods

### Cohort Definition

Using SBUH EHR data accessed through TriNetX^15^, we identified 2,466 adults aged 18 years or older with a HF diagnosis (defined by ICD-10-CM code I50) between January 1, 2016, and June 30, 2025, who newly initiated either a GLP-1 RA or an SGLT2i. Patients with an estimated glomerular filtration rate (eGFR) less than 25 mL/min/1.73 m^2^ at baseline were excluded. Individuals with documented use of both GLP-1 RA and SGLT2i were also excluded. To identify new users, we required a 12-month washout period with no prior recorded use of either treatment. The index date was defined as the date of treatment initiation. The outcome was a composite of all-cause mortality or HF-related hospitalization (defined as HF diagnosis recorded during inpatient encounter) within 1 year following the index date. Baseline demographic and clinical characteristics were summarized to describe the study population.

### Feature Selection

Clinically relevant covariates were selected a priori based on domain expertise and availability during a 12-month lookback period prior to the index date. These included demographics (sex, race, ethnicity), clinical characteristics (age at index, years from HF diagnosis to index), and pre-existing comorbidities, including atherosclerosis, coronary artery disease (CAD), chronic liver disease, chronic lung disease, chronic kidney disease (CKD), cerebrovascular disease (CVD), and T2DM. Baseline medication use included angiotensin-converting enzyme inhibitors or angiotensin receptor blockers (ACE/ARBs), BB, MRA, calcium channel blockers (CCB), statins, loop diuretics, thiazides, other anti-hypertensives, insulin, metformin, sulfonylurea, dipeptidyl peptidase-4 inhibitors (DPP4i), and thiazolidinediones (TZD). Baseline laboratory and clinical measurements (defined as closest non-missing values prior to the index date) included eGFR, creatinine, hemoglobin A1c, glucose, hemoglobin, total bilirubin, blood urea nitrogen (BUN), albumin, leukocytes (WBC), troponin, N-terminal pro-B-type natriuretic peptide (NT-proBNP), potassium, sodium, chloride, diastolic blood pressure (DBP), systolic blood pressure (SBP), and body mass index (BMI). HF phenotype was categorized as HFpEF, HFrEF, or others. Missing continuous variables were imputed using the median value. To account for the potential informativeness of missing data, indicator variables for missing covariates were included.

### Causal Inference Framework

We employed a doubly robust causal inference framework that combines nuisance models for treatment assignment and outcome risk to adjust for confounding and estimate counterfactual outcomes. This approach yields consistent treatment effect estimates if either the treatment model or the outcome model is correctly specified^16^.

#### 1. Nuisance Model Development

Two nuisance models were estimated within the doubly robust framework: a treatment assignment model and an outcome regression model^16^. Let A denote treatment assignment, where A = 1 corresponds to GLP-1 RA and A = 0 corresponds to SGLT2i. Let W denote the vector of baseline covariates, and let Y denote the binary composite outcome of all-cause mortality or HF-related hospitalization within 1 year of index. The treatment assignment model estimated the propensity score, defined as ĝ(W) = P(A = 1 | W), representing the probability of initiating GLP-1 RA conditional on baseline covariates. The outcome regression model estimated T-learner style arm-specific conditional outcome risks, defined as 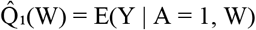 and 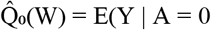, representing the expected probability of the composite outcome under each treatment given baseline covariates. All covariates described in the Feature Selection section were included in both nuisance models.

Both nuisance functions were estimated using a stacked Super Learner ensemble machine learning approach^17^. A stacked ensemble combines predictions from multiple base learners using a meta learner trained on cross-validated predictions, allowing the final model to adaptively weight different modeling approaches. In this study, the base learners included random forests, gradient-boosted trees, and L2-regularized logistic regression, and we used logistic regression to construct the final ensemble (the meta learner)^17^. Five-fold stratified cross-validation was used within the stacking procedure to generate out-of-fold predicted probabilities for training the meta learner, reducing overfitting and improving robustness of nuisance function estimation.

#### 2. Nuisance Model Diagnostics

Diagnostic assessments were conducted to evaluate whether the nuisance models were adequate for supporting stable estimation of treatment effects. For the treatment assignment model, overlap of estimated propensity scores between treatment groups was examined to assess the plausibility of the positivity assumption^12^. Calibration of the treatment model was evaluated by comparing observed treatment proportions with mean predicted probabilities across propensity score deciles. Covariate balance between treatment groups was assessed using standardized mean differences (SMD) before and after inverse probability of treatment weighting (IPTW)^18^. IPTW creates a pseudo-population in which baseline covariates are independent of treatment assignment, approximating the balance that would be achieved through randomization. SMD <0.10 indicates an acceptable balance between the two groups.

For the outcome regression model, discriminative performance was evaluated using the area under the receiver operating characteristic curve (AUC-ROC) based on out-of-fold predicted risks for the observed treatment assignment, both overall and within each treatment arm. Model calibration and overall predictive performance were further evaluated using the Brier score, log loss, calibration slope, and calibration intercept. To improve transparency, permutation-based feature importance was used to identify predictors that contributed most to the outcome models.

#### 3. Average Treatment Effect (ATE) Estimation

The ATE is a population-level causal estimand that quantifies the difference in expected outcomes if all individuals received one treatment versus the other^9^. Formally, the ATE (risk difference) is defined as ATE = E[Y(1) − Y(0)], where Y(1) and Y(0) denote the potential outcomes under initiation of a GLP-1 RA and an SGLT2i, respectively. The primary ATE was estimated using a doubly robust augmented inverse probability weighting (AIPW) estimator^16^. This approach combines information from both the treatment assignment model and the outcome regression model and yields consistent estimates if either nuisance model is correctly specified.

The AIPW estimator is given 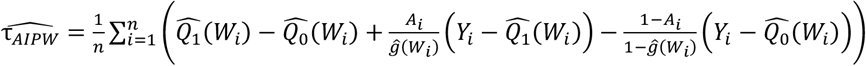 where ĝ(W_i_) is the estimated propensity score, and 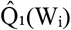 and 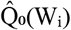 are the estimated arm-specific outcomes. In addition to ATE, risk ratios (RR) were calculated as RR = E[Y(1)] / E[Y(0)] to provide a relative measure of treatment effect. For baseline comparison, a crude unadjusted ATE estimate was calculated as the difference in observed outcome rates between treatment groups, ignoring covariates. In addition, an adjusted outcome regression model was fit using logistic regression with treatment and baseline covariates as predictors.

#### 4. Sensitivity and Robustness Analyses

To assess sensitivity to potential unmeasured confounding, the E-value was computed for the estimated ATE^19^. As sensitivity analyses, targeted maximum likelihood estimation (TMLE)^20^ and IPTW with stabilized weights^18^ were used to estimate the ATE and evaluate the consistency of results across alternative causal estimators. Trimming sensitivity analyses were conducted by restricting the cohort to regions where propensity scores of GLP-1 RA and SGLT2i overlap (0.05 ≤ ĝ ≤ 0.95, 0.10 ≤ ĝ ≤ 0.90), then re-estimating the ATE using AIPW within each trimmed subset^21^. Two robustness checks were performed, treatment permutation and random noise covariate addition, to assess the susceptibility of the findings to spurious associations^12^.

#### 5. Subgroup Analyses

Clinical subgroup analysis was conducted by estimating the AIPW-ATE within strata defined a priori based on HF-relevant baseline characteristics. These included age, sex, BMI, eGFR, comorbidity of CAD and T2DM, loop diuretic use, and HF phenotype. Subgroup-specific ATE estimates were summarized using a forest plot to facilitate comparison across subgroups. In addition, heterogeneity analysis was performed by testing differences in treatment effects between subgroup levels to assess potential effect modification^22^. To account for multiple comparisons across the prespecified heterogeneity tests, we controlled the false discovery rate (FDR) using the Benjamini– Hochberg procedure and reported both nominal and FDR-adjusted p values^23^.

#### 6. Conditional Average Treatment Effect (CATE) Estimation

The CATE is an individual-level causal estimand that characterizes treatment effect heterogeneity by quantifying how the expected treatment effect varies with baseline covariates^9^. Formally, the CATE is defined as CATE(x)=E[Y(1)−Y(0)∣X=x], where X denotes a vector of individual-level baseline covariates. CATE estimation was conducted using the doubly robust learner (DRLearner)^24^ implemented in the econml Python package as the primary approach^25^. In this framework, estimates from the treatment assignment and outcome regression nuisance models were combined to construct doubly robust pseudo-outcomes derived from the AIPW estimating equation. These pseudo-outcomes were then regressed on baseline covariates using a random forest regressor as the final stage model to estimate individual treatment effects. As a sensitivity analysis, causal forest with double machine learning (CausalForestDML)^26,27^, also implemented in econml, was applied to flexibly model potential treatment effect heterogeneity while orthogonalizing for nuisance parameters, providing an alternative nonparametric approach to CATE estimation.

#### 7. CATE Analyses

Several exploratory analyses were conducted to characterize and evaluate estimated CATEs. The distribution of individual-level CATE estimates was examined to summarize the magnitude and variability of heterogeneity. As a sensitivity analysis, we restricted the cohort to regions of propensity score overlap (0.05 ≤ ĝ ≤ 0.95, 0.10 ≤ ĝ ≤ 0.90), then re-estimated CATE values by refitting the DRLearner procedure within each trimmed subset^21^. Calibration of CATE estimates was assessed by stratifying individuals into deciles based on predicted CATE values and comparing AIPW-estimated effects across strata^28^. An individualized treatment policy that assigned treatment based on patient-level CATE estimates was evaluated to assess potential decision-relevant implications^29^. Permutation-based feature importance was examined to identify baseline covariates associated with variation in estimated treatment effects. Baseline characteristics were summarized across CATE quartiles. All code for analyses and diagnostic results is available in the project GitHub repository (https://github.com/gracehan0706/HF-Causal-ML).

## Results

### Cohort Characteristics

The study cohort included 2,466 adults with HF who initiated either a GLP-1 RA (n=567) or SGLT2i (n=1,899). Overall, patients were mostly male (62%) and predominantly white (78%). HF phenotype differed between groups, with GLP-1 RA initiators more likely to have HFpEF and SGLT2i initiators more likely to have HFrEF. GLP-1 RA initiators were slightly younger (mean age: 67.4 years), initiated therapy later after HF diagnosis (2.1 years), and had worse cardiometabolic risk profiles, including higher prevalence of obesity and T2DM, higher BMI, and higher baseline HbA1c. In contrast, SGLT2i initiators were slightly older (mean age: 68.9 years), initiated therapy sooner after HF diagnosis (1.4 years), and more frequently used cardiovascular medications, including ACE/ARBs, BB, MRA, CCB, and thiazide diuretics. During 1 year of follow up, the composite outcome occurred in 453 of 2,466 patients (18.45%).

### Nuisance Model Diagnostics

Estimated propensity scores ranged from 0.042 to 0.948 with reasonable overlap between treatment groups. However, a meaningful fraction had low propensity for GLP-1 RA initiation (<0.05: 9.5%; <0.10: 43.8%) (Figure 1A). Extreme high propensities were uncommon (>0.90: 0.77%; >0.95: 0%). IPTW effective sample sizes were 315 of 567 for the treated group and 1,410 of 1,899 for the control group, suggesting moderate weight variability without domination by a small number of observations. Decile-based calibration demonstrated close agreement between observed and predicted treatment probabilities (Figure 1B). Discrimination was high (AUC 0.855), and overall prediction error was low (Brier score 0.127), consistent with the observed decile-based calibration. Before inverse propensity treatment weighting (IPTW), covariate imbalance was substantial, with a mean absolute SMD of 0.224 (maximum 0.816) and 43 of 64 covariates exceeding 0.1. After IPTW, balance improved meaningfully, with the mean absolute SMD decreasing to 0.089 (maximum 0.344) and 15 covariates remaining above 0.1 (T2DM, HbA1c, insulin, BMI, metformin, MRA, troponin_missing, NT-proBNP, glucose, NT-proBNP_missing, albumin, BMI_missing, sulfonylurea, bilirubin_missing, and HFrEF). Weight diagnostics suggested moderate tail behavior (unstabilized IPTW: median 1.14, 90^th^ 3.20, 95^th^ 5.27, 99^th^ 11.17, max 19.20). As a sensitivity analysis, stabilized IPTW weights were examined and were less variable (median 0.84, 90^th^ 1.44, 95^th^ 2.01, 99^th^ 3.82, max 10.15), yielding similar balance diagnostics.

**Figure 1.**
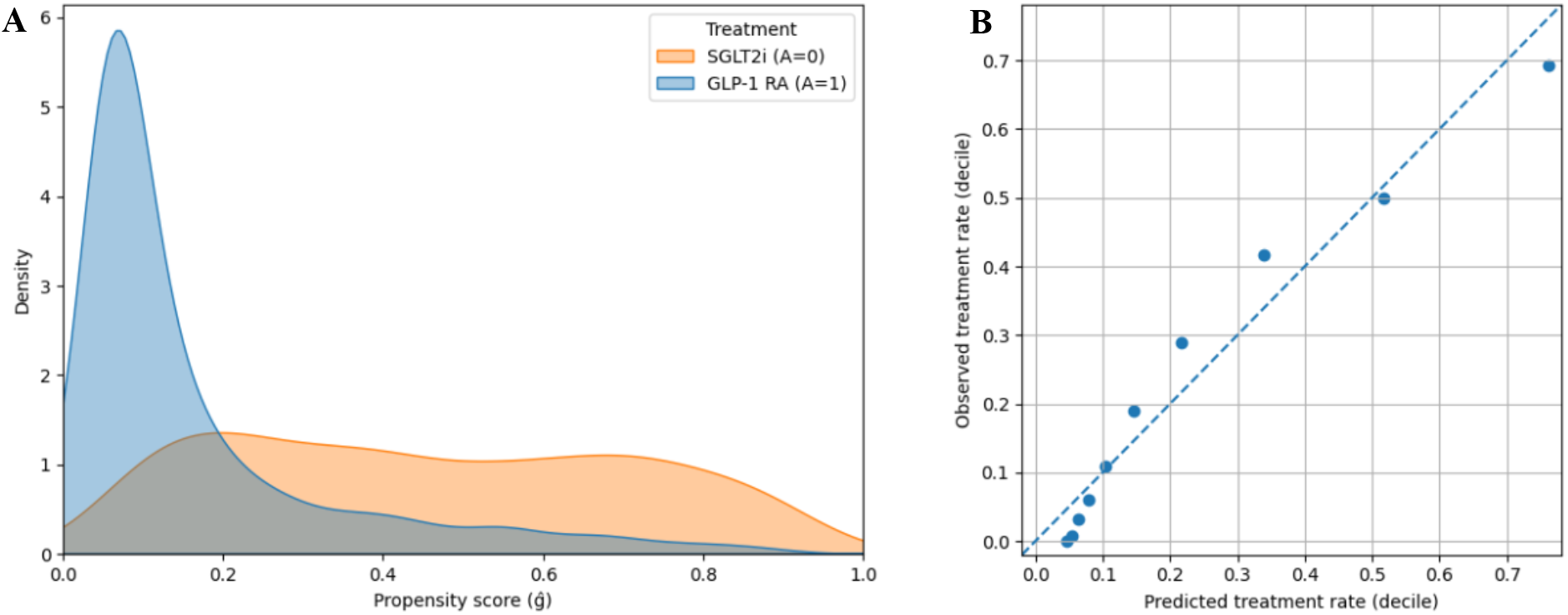
Propensity score diagnostics. (**A**) Kernel density distribution of estimated propensity scores by treatment group: SGLT2i (A=0) and GLP-1 RA (A=1). (**B**) Decile-based calibration plot of observed versus predicted treatment probabilities across propensity score deciles. The dashed line represents perfect calibration.

For the outcome model, out-of-fold diagnostic evaluation demonstrated moderate overall discriminative performance (AUC 0.692). Discrimination within the SGLT2i arm was similar (AUC 0.672), whereas discrimination within the GLP-1 RA arm was limited (AUC 0.463), likely reflecting the smaller sample size and lower event rate in that subgroup. Calibration metrics remained acceptable, with an overall Brier score of 0.139, log loss of 0.443, calibration intercept of 0.049, and calibration slope of 1.038. The top predictive features for the arm-specific outcome models largely align with established clinical prognostic markers. Detailed feature importance results are available in the project GitHub repository. Taken together, these diagnostics suggest that the nuisance models generated stable probability estimates for use in downstream causal analyses.

### ATE Estimation

The composite outcome occurred in 8.11% of GLP-1 RA initiators and 21.43% of SGLT2i initiators. Under the primary AIPW analysis, the estimated 1-year composite outcome risk was 10.1% under universal GLP-1 RA initiation and 19.2% under universal SGLT2i initiation. This resulted in a risk difference (ATE) of −0.091 (95% CI −0.127 to −0.056), corresponding to a 9.1% lower risk of 1-year all-cause mortality or HF-hospitalization under GLP-1 RA initiation compared with SGLT2i initiation. A risk ratio (RR) of 0.524 (95% CI 0.381 to 0.720) indicated a 47.4% lower risk of the composite outcome under GLP-1 RA initiation (Table 1).

**Table 1.** Average treatment effect (ATE) estimates and corresponding risk ratios (RR) for GLP-1 RA versus SGLT2i across causal estimators.

| Category | Estimator | ATE | 95% CI | RR | 95% CI |
| --- | --- | --- | --- | --- | --- |
| Baseline | Unadjusted | -0.133 | [-0.162, -0.104] | 0.379 | [0.266, 0.493] |
|  | Adjusted outcome regression | -0.073 | [-0.111, -0.036] | 0.630 | [0.457, 0.832] |
| Doubly Robust | AIPW | -0.091 | [-0.127, -0.056] | 0.524 | [0.381, 0.720] |
| Sensitivity Analysis | TMLE | -0.075 | [-0.110, -0.039] | 0.616 | [0.470, 0.807] |
|  | IPTW | -0.088 | [-0.129, -0.046] | 0.542 | [0.353, 0.752] |
| | AIPW ($0.05 \leq \hat{g} \leq 0.95$) | -0.083 | [-0.121, -0.045] | 0.553 | [0.395, 0.776] |
| | AIPW ($0.10 \leq \hat{g} \leq 0.90$) | -0.062 | [-0.098, -0.025] | 0.590 | [0.416, 0.837] |

Abbreviations: AIPW, augmented inverse propensity weighting; TMLE, targeted maximum likelihood estimation; IPTW, inverse probability of treatment weighting.

### Sensitivity and Robustness Analyses

The E-value for the primary ATE estimate was 3.227, indicating that an unmeasured confounder would need to be associated with both treatment assignment and the outcome by a risk ratio of at least 3.227 each to fully explain away the estimated treatment effect. Sensitivity analyses using TMLE and stabilized IPTW produced similar estimates (Table 1), demonstrating consistency across causal estimators. Propensity score trimming to 0.05 ≤ ĝ ≤ 0.95 retained 2,231 of 2,466 patients (90.5%) and yielded an AIPW-estimated ATE similar to that of the full cohort. A more restrictive trim to 0.10 ≤ ĝ ≤ 0.90 retained 1,367 patients (55.4%) and produced an attenuated ATE that still favored GLP-1 RA initiation.

Two robustness checks were performed to evaluate the stability of the estimated treatment effect. Under treatment permutation, the mean estimated ATE was 0.004 (SD 0.022). This is consistent with the null hypothesis of no causal effect, indicating that the estimator did not exhibit systematic bias under randomized treatment assignment. In the random noise covariate addition test, the mean ATE was −0.093 (SD 0.003), closely aligned with the primary estimate. These findings suggest that the AIPW-estimated ATE was not influenced by model instability or sensitivity to irrelevant covariates.

### Subgroup Analyses

Subgroup-specific ATE estimates were directionally consistent with the overall treatment effect across predefined demographic and clinical strata (Figure 2), suggesting a protective association for GLP-1 RA initiation. Although confidence intervals were wider in smaller subgroups, most subgroup estimates overlapped with the overall ATE, and the magnitude of effect appeared broadly similar across subgroups.

**Figure 2.**
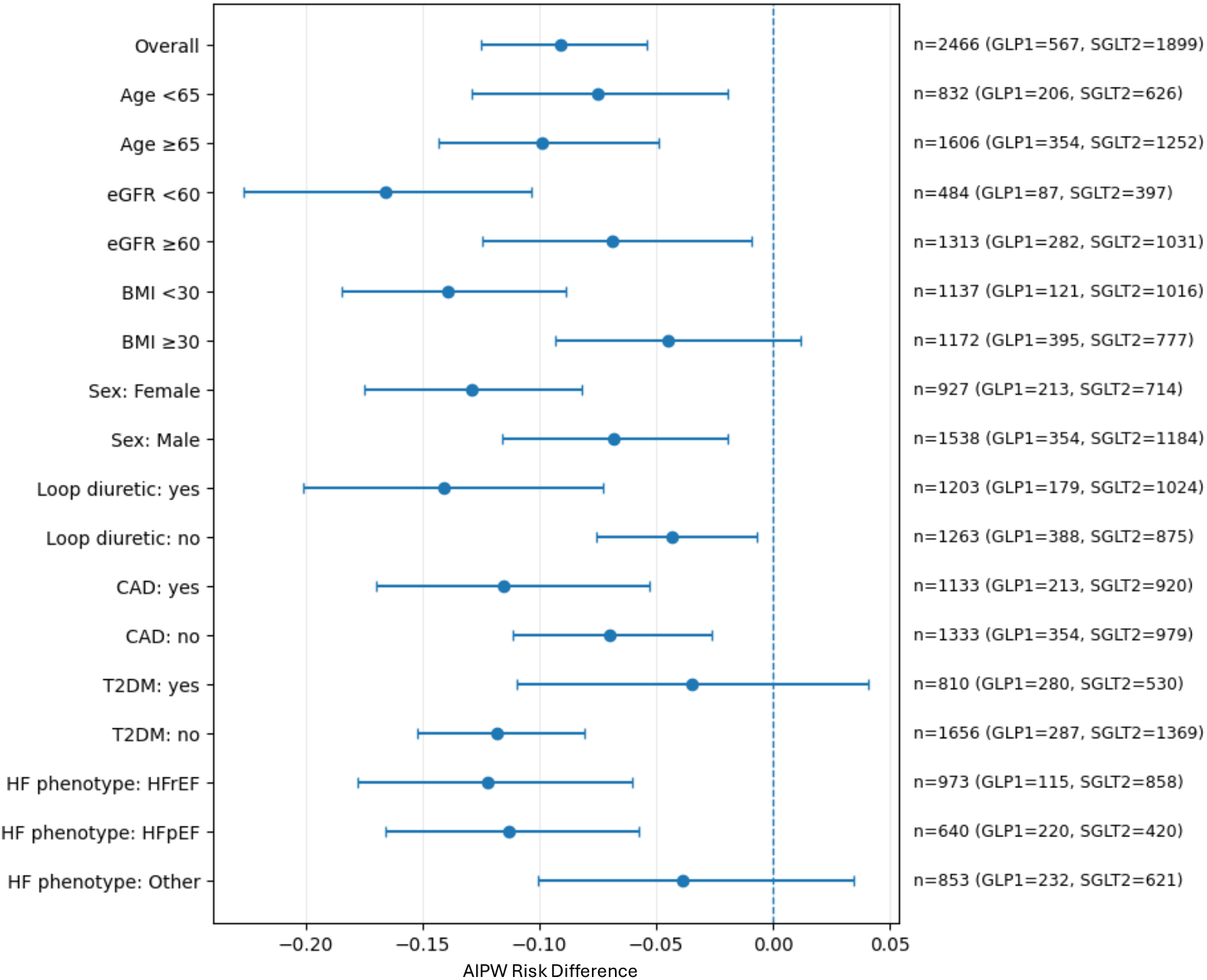
Forest plot of subgroup-specific risk differences. Points represent subgroup-specific average treatment effects estimated using augmented inverse propensity weighting (AIPW), and horizontal lines indicate 95% confidence intervals. The vertical dashed line denotes no difference. Sample sizes are shown on the right.

Subgroup interaction tests using bootstrap-based comparisons of doubly robust pseudo-outcomes identified evidence of effect modification by loop diuretic use, BMI, and eGFR. Patients receiving loop diuretics, those with reduced kidney function (eGFR <60 mL/min/1.73 m^2^), those with a BMI <30, and those without a comorbidity of T2DM experienced larger risk reductions with GLP-1 RA (nominal p = 0.013, 0.014, 0.020, 0.042, respectively). After controlling for the FDR using the Benjamini-Hochberg procedure, loop diuretic use, eGFR, and BMI remained significant (adjusted p = 0.040 for each), whereas T2DM did not (adjusted p = 0.063).

### CATE Analyses

To evaluate potential treatment effect heterogeneity, CATEs were estimated using DRLearner as the primary approach. Individual-level CATE estimates were centered near the overall ATE (mean −0.067, SD 0.114), indicating modest dispersion in predicted treatment effects (Figure 3A). Propensity score trimming yielded similar CATE distributions (0.05 ≤ ĝ ≤ 0.95: mean −0.069, SD 0.118; 0.10 ≤ ĝ ≤ 0.90: mean −0.057, SD 0.110), suggesting that extreme propensity regions did not drive the ITE patterns. Calibration of predicted CATE estimates demonstrated limited agreement between observed and predicted treatment effects across deciles (Figure 3B). Observed AIPW-estimated effects showed substantial overlap across strata with wide confidence intervals, implying instability in predicted heterogeneity.

**Figure 3.**
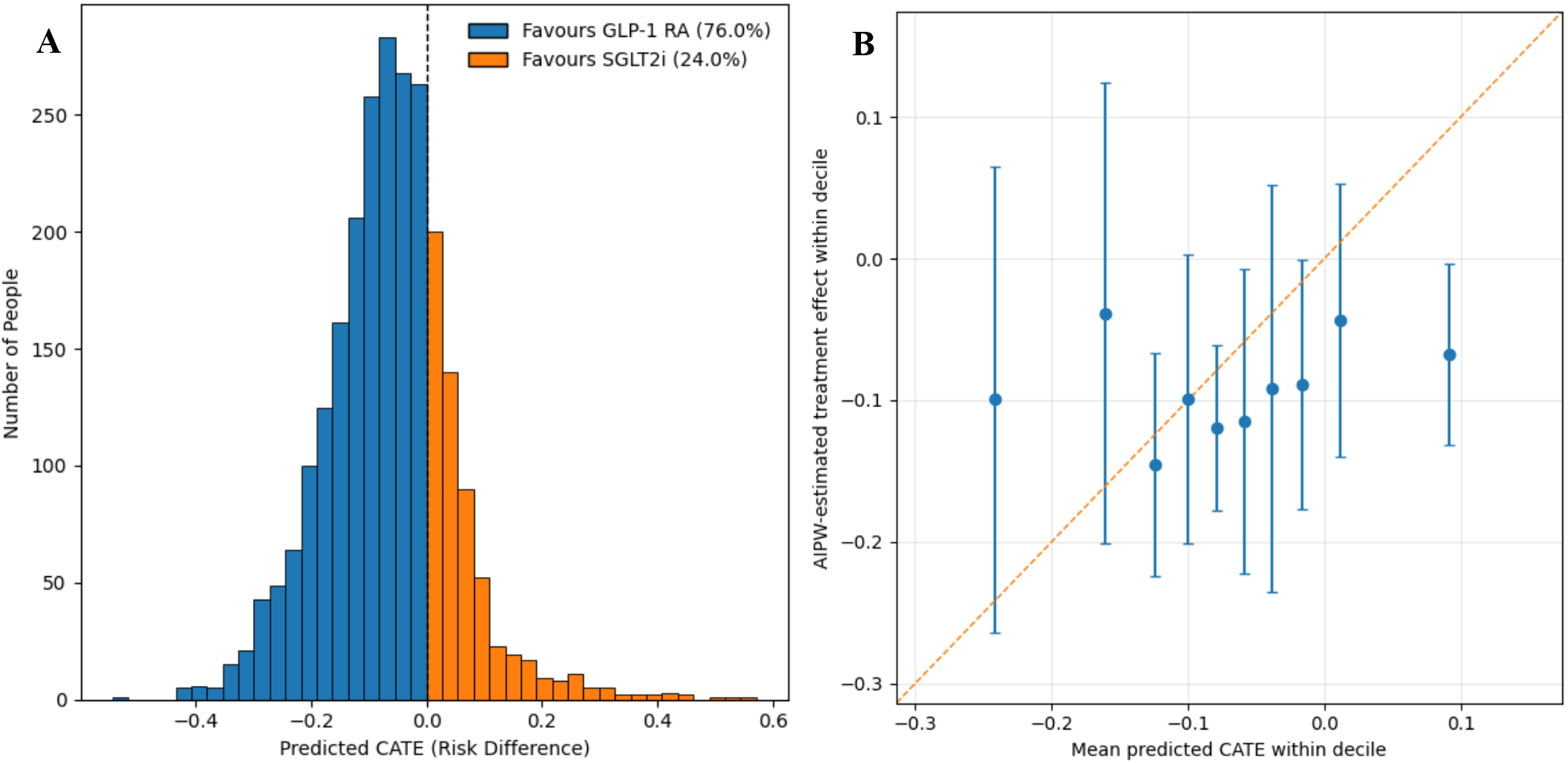
Predicted conditional average treatment effects (CATE) and calibration of DRLearner. (**A**) Distribution of CATE estimates for GLP-1 RA versus SGLT2i. Negative values indicate a predicted benefit for GLP-1 RA, whereas positive values indicate a predicted benefit for SGLT2i. (**B**) Decile-based calibration plot of AIPW-estimated treatment effects with mean predicted CATE estimates. The orange diagonal line indicates perfect calibration.

An individualized treatment policy derived from predicted CATE estimates did not improve expected outcomes compared with assigning all individuals to GLP-1 RA. The estimated policy value, defined as the expected outcome risk under the treatment assignment rule, was 0.112 (95% CI 0.082 to 0.143). This was higher than the value under universal GLP-1 RA treatment (0.101, 95% CI 0.070 to 0.132) but lower than universal SGLT2i treatment (0.192, 95% CI 0.173 to 0.210). These findings suggest that model-based heterogeneity did not translate into a clinically beneficial individualized treatment strategy beyond the overall population-level effect.

Permutation-based feature importance was used to assess baseline characteristics contributing to variability in predicted CATE estimates. Loop diuretic use demonstrated the highest relative importance (16.1%), followed by NT-proBNP (15.1%), HbA1c (13.4%), BUN (10.5%), and WBC (8.6%). Additional variables, including BMI, DBP, glucose, age at index, and hemoglobin, contributed more modestly to predicted heterogeneity. Importance was distributed across multiple covariates rather than concentrated in a single dominant feature. Consistent with CATE calibration and individualized policy evaluations, these findings indicate limited heterogeneity in treatment effects.

CausalForestDML was implemented as a sensitivity analysis and yielded results consistent with DRLearner, without evidence of robust or clinically actionable heterogeneity. Overall, model-based CATE analyses suggest limited treatment effect variation beyond the average population-level effect. All CausalForestDML results are available in the project GitHub repository.

## Discussion

In this EHR-based comparative effectiveness study, we estimated a population-level ATE favoring the initiation of GLP-1 RA over SGLT2i for lowering the 1-year risk of the composite outcome under a doubly robust causal ML framework. However, we did not observe convincing or clinically coherent CATE patterns. Causal ML aims to predict counterfactual outcomes, namely, what a patient’s outcome would have been under GLP-1 RA versus under SGLT2i initiation. Since only one of these potential outcomes is observed for each patient, treatment effects are not directly identifiable without assumptions about the data-generating process. Therefore, we interpret our findings under three standard identification assumptions in causal inference: unconfoundedness (ignorability), positivity (overlap), and the stable unit treatment value assumption (SUTVA)^12^.

First, unconfoundedness requires that treatment initiation be independent of potential outcomes conditional on measured baseline covariates. In this cohort, however, confounding by indication was substantial. Clinically, GLP-1 RA is often initiated among patients with higher BMI and worse glycemic control, whereas SGLT2i is more commonly initiated among patients with higher cardiovascular risk and greater illness severity. Consistent with this, event rates were markedly higher among SGLT2i initiators than GLP-1 RA initiators (21.43% vs. 8.11%). When higher risk patients preferentially receive SGLT2i, CATE models may attribute a larger counterfactual benefit to GLP-1 RA, even in the absence of true pharmacologic effect modification. The BMI-stratified results illustrate this mechanism. Among SGLT2i initiators, event rates were higher for BMI <30 than BMI ≥30 (24.2% vs. 17.8%), while GLP-1 RA initiator event rates were similar across BMI strata (about 8.3%), producing a larger risk difference in BMI <30 (−13.5%) than BMI ≥30 (−4.8%). Although a larger apparent GLP-1 RA benefit in BMI <30 may seem counterintuitive, BMI <30 in HF can reflect frailty or more advanced disease, indicating a higher baseline risk. Similarly, patients predicted to benefit most from GLP-1 RA had higher NT-proBNP levels, greater loop diuretic use, and higher use of concomitant cardiometabolic medications and HF-related therapies, all markers of greater disease burden and baseline risk. Since key severity determinants are incompletely captured in EHR data, unconfoundedness cannot be verified, and residual confounding remains plausible.

A related challenge is workflow-dependent measurement. Several of the largest separations across CATE strata involved laboratory missingness indicators such as albumin, troponin, NT-proBNP, glucose, and hemoglobin. In routine practice, these tests are ordered based on acuity and setting. For example, troponin and NT-proBNP are often obtained during acute hospitalizations. As a result, whether a lab is recorded or not serves as a proxy for severity and encounter context. When heterogeneity models leverage these workflow signals to stratify patients, they can generate apparent CATE variation that reflects the clinical reality of data generation.

Second, positivity requires that both treatments are plausible options across the baseline covariate profiles. The fraction of individuals with a propensity score near 0 suggested that GLP-1 RA initiation was rare for certain covariate profiles. This imposes practical positivity limits, such that counterfactual predictions in these regions may rely more heavily on extrapolation. However, a mild propensity score trim produced minimal changes in ATE and CATE, whereas a more restrictive trim attenuated the estimated effects but remained directionally consistent. These results suggest that overlap limitations are unlikely to explain the overall beneficial effect on GLP-1 RA initiation. Nevertheless, they may influence its magnitude and further constrain the stability and clinical interpretability of heterogeneity patterns under real-world prescribing.

Third, SUTVA (consistency) requires well-defined treatment strategies and no interference between patients. Interference is likely minimal here, as one patient’s GLP-1 RA or SGLT2i initiation is unlikely to affect another patient’s outcome. Consistency is more challenging because initiation can represent heterogeneous treatment versions such as agent choice, dosing, adherence, discontinuation, and switching. Without modeling these post-initiation dynamics, some variability in model estimates may reflect differences in treatment versions and care trajectories rather than true heterogeneity.

Although we used a doubly robust AIPW framework with multiple sensitivity and robustness analyses, valid inference still depends on adequate covariate overlap and the absence of important unmeasured confounding, both of which are inherently challenging in real-world EHR studies. In our data, IPTW weighting improved covariate balance, but residual differences persisted in cardiometabolic, HF severity-related, and missingness indicators. Even though subgroup heterogeneity tests remained significant for eGFR, loop diuretic use, and BMI after FDR adjustment, these factors likely reflect baseline risk gradients rather than true effect modifiers, given weak CATE calibration. Nevertheless, the estimated population-level ATE remained robust, with the direction and magnitude broadly consistent across estimators and under overlap-restricted propensity trimming. Importantly, numerical stability across causal estimators should not be interpreted as proof of a causal or clinically actionable effect, because stable bias can persist across methods when key identification conditions are only partially met.

The cardiometabolic phenotype of our cohort, which was predominantly obese (mean BMI 31, IQR 26–35), provides biological plausibility for an ATE favoring GLP-1 RA initiation in the real-world setting. This interpretation is supported by emerging evidence in obesity-related HFpEF populations, as seen in STEP-HFpEF and SUMMIT^6,7^. Additional indirect support comes from a US claims-based study in which GLP-1 RA was associated with more than 40% lower risk of HF hospitalization or all-cause mortality compared with a placebo proxy (sitagliptin, a DPP4i)^30^. However, direct HF-specific head-to-head evidence remains limited and mixed. A TriNetX propensity-matched cohort of patients with HFpEF and DM reported superior cardiovascular protective effects of GLP-1 RA compared with SGLT2i^31^. Meanwhile, a Medicare cohort of patients with HFrEF and HFpEF found lower rates of HF hospitalization with SGLT2i than with GLP-1 RA^32^. Taken together, our observed ATE should be viewed as a real-world association shaped by cohort phenotype and prescribing structure, rather than as definitive evidence that GLP-1 RA outperforms SGLT2i for HF outcomes.

From a learning health system perspective, these findings highlight both the opportunity and the constraints of EHR-based causal modeling. Real-world data enable scalable comparative effectiveness analyses in populations often underrepresented in trials, including patients with multimorbidity and high obesity prevalence. However, when prescribing is structured by baseline risk, observational models can partially conflate disease severity and care pathways with treatment effects. ITE estimates derived from such data may therefore reproduce historical allocation patterns rather than reveal true biological heterogeneity. Before translating these models into clinical decision support, careful evaluation is essential to ensure transportability and clinical safety, including external and temporal validation, restriction to overlap-supported populations, and prospective silent-mode testing. If individualized policies remain well calibrated and beneficial under these safeguards, they could support patient-specific, evidence-based treatment selection, translating real-world observational data into actionable precision care.

## Conclusion

In this EHR-based comparative effectiveness study, a doubly robust causal ML framework identified a stable population-level benefit favoring the initiation of GLP-1 RA over SGLT2i in patients with HF for reducing the 1-year risk of all-cause mortality or HF-related hospitalization. However, heterogeneity analyses did not identify robust or clinically meaningful patterns to support individualized treatment selection. These findings support the use of EHR-based causal ML for population-level comparative effectiveness estimation, while underscoring caution in extending such models to patient-level treatment recommendations.

## Data Availability

All data produced in the present study are available upon reasonable request to the authors

## References

1. Bozkurt B, Ahmad T, Alexander K, et al. HF STATS 2024: Heart failure epidemiology and outcomes statistics. J Card Fail 2025; 31: 66–116.

2. Simmonds SJ, Cuijpers I, Heymans S, et al. Cellular and molecular differences between HFpEF and HFrEF: a step ahead in an improved pathological understanding. Cells 2020; 9: 242.

3. Shahid I, Khan MS, Butler J, et al. Initiation and sequencing of guideline-directed medical therapy for heart failure across the ejection fraction spectrum. Heart Fail Rev 2025; 30: 515–523.

4. Vaduganathan M, Docherty KF, Claggett BL, et al. SGLT2 inhibitors in patients with heart failure: a comprehensive meta-analysis of five randomised controlled trials. Lancet 2022; 400: 757–767.

5. Pierce JB, Vaduganathan M, Fonarow GC, et al. Contemporary use of sodium-glucose cotransporter-2 inhibitor therapy among patients hospitalized for heart failure with reduced ejection fraction in the US: the Get With The Guidelines-Heart Failure Registry. JAMA Cardiol 2023; 8: 652–661.

6. Rivera FB, Cruz LLA, Magalong JV, et al. Cardiovascular and renal outcomes of glucagon-like peptide 1 receptor agonists among patients with and without type 2 diabetes mellitus: a meta-analysis of randomized placebo-controlled trials. Am J Prev Cardiol 2024; 18: 100679.

7. Rahmani AR, Dhaliwal SK, Pastena P, et al. GLP-1 receptor agonists in heart failure. Biomolecules 2025; 15: 1403.

8. Kent DM, Rothwell PM, Ioannidis JP, et al. Assessing and reporting heterogeneity in treatment effects in clinical trials: a proposal. Trials 2010; 11: 85.

9. Rubin DB. Estimating causal effects of treatments in randomized and nonrandomized studies. J Educ Psychol 1974; 66: 688–701.

10. Hernán MA, Robins JM. Using big data to emulate a target trial when a randomized trial is not available. Am J Epidemiol 2016; 183: 758–764.

11. Stuart EA, DuGof E, Abrams M, et al. Estimating causal effects in observational studies using electronic health data: challenges and (some) solutions. EGEMs 2013; 1: 4.

12. Feuerriegel S, Frauen D, Melnychuk V, et al. Causal machine learning for predicting treatment outcomes. Nat Med 2024; 30: 958–968.

13. Wang T, Pate V, Wyss R, et al. High-dimensional iterative causal forest (hdiCF) for subgroup identification using health care claims data. Am J Epidemiol 2024; 194: 2085–2097.

14. Cardoso P, Young KG, Nair ATN, et al. Phenotype-based targeted treatment of SGLT2 inhibitors and GLP-1 receptor agonists in type 2 diabetes. Diabetologia 2024; 67: 822–836.

15. Palchuk MB, London JW, Perez-Rey D, et al. A global federated real-world data and analytics platform for research. JAMIA Open 2023; 6: ooad035.

16. Glynn AN, Quinn KM. An introduction to the augmented inverse propensity weighted estimator. Polit Anal 2010; 18: 36–56.

17. Butzin-Dozier Z, Ji Y, Coyle J, et al. Treatment heterogeneity of water, sanitation, hygiene, and nutrition interventions on child growth by environmental enteric dysfunction and pathogen status for young children in Bangladesh. PLoS Negl Trop Dis 2025; 19: e0012881.

18. Austin PC, Stuart EA. Moving towards best practice when using inverse probability of treatment weighting (IPTW) using the propensity score to estimate causal treatment effects in observational studies. Stat Med 2015; 34: 3661–3679.

19. VanderWeele TJ, Ding P. Sensitivity analysis in observational research: introducing the E-value. Ann Intern Med 2017; 167: 268–274.

20. Levy J, van der Laan M, Hubbard A, Pirracchio R. A fundamental measure of treatment effect heterogeneity. J Causal Inference 2021; 9: 83–108.

21. Petersen ML, Porter KE, Gruber S, et al. Diagnosing and responding to violations in the positivity assumption. Stat Methods Med Res 2012; 21: 31–54.

22. Brookes ST, Whitely E, Egger M, et al. Subgroup analyses in randomized trials: risks of subgroup-specific analyses. J Clin Epidemiol 2004; 57: 229–236.

23. Benjamini Y, Hochberg Y. Controlling the false discovery rate: a practical and powerful approach to multiple testing. J R Stat Soc Ser B Stat Methodol 1995; 57: 289–300.

24. Kennedy EH. Towards optimal doubly robust estimation of heterogeneous causal effects. Electron J Stat 2023;17(2):3008–3049.

25. Battocchi K, Dillon E, Hei M, Lewis G, Oka P, Oprescu M, Syrgkanis V. EconML: a Python package for ML-based heterogeneous treatment effects estimation. 2019.

26. Wager S, Athey S. Estimation and inference of heterogeneous treatment effects using random forests. J Am Stat Assoc 2018;113(523):1228–1242.

27. Chernozhukov V, Chetverikov D, Demirer M, et al. Double/debiased machine learning for treatment and structural parameters. Econom J 2018; 21: C1–C68.

28. Chernozhukov V, Demirer M, Duflo E, et al. Fisher–Schultz lecture: generic machine learning inference on heterogeneous treatment effects in randomized experiments, with an application to immunization in India. Econometrica 2025; 93: 1121–1164.

29. Dudík M, Erhan D, Langford J, et al. Doubly robust policy evaluation and optimization. Stat Sci 2014;29(4):485–511.

30. Krüger N, Schneeweiss S, Fuse K, et al. Semaglutide and tirzepatide in patients with heart failure with preserved ejection fraction. JAMA 2025; 334(14): 1255–1266.

31. Li AC-W, Lin Y-C, Huang J-Y, et al. Superior cardiovascular protection with GLP-1 RAs over SGLT2 inhibitors in DM and HFpEF: a propensity score matching study. PLoS One 2025; 20: e0326534.

32. Gonzalez J, Bates BA, Setoguchi S, et al. Cardiovascular outcomes with SGLT2 inhibitors versus DPP4 inhibitors and GLP-1 receptor agonists in patients with heart failure with reduced and preserved ejection fraction. Cardiovasc Diabetol 2023; 22: 54.

